# Acute ischemic stroke alters the brain’s preference for distinct dynamic connectivity states

**DOI:** 10.1101/19011031

**Authors:** Anna K. Bonkhoff, Flor A. Espinoza, Harshvardhan Gazula, Victor M. Vergara, Lukas Hensel, Jochen Michely, Theresa Paul, Anne Rehme, Lukas J. Volz, Gereon R. Fink, Vince D. Calhoun, Christian Grefkes

**Author notes:** corresponding author: Christian Grefkes, Department of Neurology, University Hospital Cologne, Cologne, Germany.

## Abstract

Acute ischemic stroke disturbs healthy brain organization, prompting subsequent plasticity and reorganization to compensate for loss of specialized neural tissue and function. *Static* resting-state functional magnetic resonance imaging (fMRI) studies have already furthered our understanding of cerebral reorganization by estimating stroke-induced changes in network connectivity aggregated over the duration of several minutes. In this study, we utilized *dynamic* resting-state fMRI analyses to increase temporal resolution to seconds and explore transient configurations of motor network connectivity in acute stroke. To this end, we collected resting-state fMRI data of 31 acute ischemic stroke patients and 17 age-matched healthy controls. Stroke patients presented with moderate to severe hand motor deficits. By estimating dynamic connectivity within a sliding window framework, we identified three distinct connectivity configurations of motor-related networks. Motor networks were organized into three regional domains, i.e. a cortical, subcortical and cerebellar domain. Temporal connectivity patterns of stroke patients markedly diverged from those of healthy controls depending on the severity of the initial motor impairment. Moderately affected patients (n=18) spent significantly more time in a weakly connected configuration that was characterized by low levels of connectivity, both locally as well as between distant regions. In contrast, severely affected patients (n=13) showed a significant preference for transitions into a spatially segregated connectivity configuration. This configuration featured particularly high levels of local connectivity within the three regional domains as well as anti-correlated connectivity between distant networks across domains. A third connectivity configuration represented an intermediate connectivity pattern compared to the preceding two, and predominantly encompassed decreased inter-hemispheric connectivity between cortical motor networks independent of individual deficit severity. Alterations within this third configuration thus closely resembled previously reported ones originating from *static* resting-state fMRI studies post-stroke.

In summary, acute ischemic stroke not only prompted changes in connectivity between distinct functional networks, yet also caused severe aberrations in temporal properties of large-scale network interactions depending on the individual deficit severity. These findings offer new vistas on the dynamic neural mechanisms underlying acute neurological symptoms, cortical reorganization and treatment effects in stroke patients.

## Introduction

Ischemic stroke is a major cause of sudden focal brain damage, which severely disrupts structural and functional integrity at both local and global scales (von Monakow, 1914; Carrera and Tononi, 2014). Functional neuroimaging has strongly contributed to reveal neural mechanisms engaged in post-stroke plasticity and reorganization (Grefkes and Fink, 2011; Grefkes and Fink, 2014; Ward, 2017). More precisely, resting-state functional magnetic resonance imaging (fMRI) studies have frequently demonstrated disturbances in inter-hemispheric connectivity following stroke (Carter *et al*., 2010; Wang *et al*., 2010; Golestani *et al*., 2013; Rehme *et al*., 2014). A highly consistent finding encountered in motor stroke is the reduction of interhemispheric connectivity between the primary sensorimotor cortices, which develops in the first weeks after stroke and returns to levels observed in healthy subjects in parallel to behavioral recovery (van Meer *et al*., 2010; Park *et al*., 2011; Volz et al., 2016). However, since conventionally applied analysis tools do not allow for a fine-grained temporal evaluation of resting-state fMRI signals, it is currently unknown whether such stroke-induced alterations in functional network connectivity additionally exhibit symptom-severity dependent fluctuations. These temporal variations could reflect network flexibility necessary for neural reorganization underlying recovery of function.

Importantly, the time resolution of resting-state fMRI data has recently been increased by the advent of time-varying or *dynamic* functional network connectivity (dFNC) analyses (Chang and Glover, 2010; Allen *et al*., 2014; Calhoun *et al*., 2014). In contrast to the previous assumption of *static* connectivity over the entire duration of an fMRI scan, *dynamic* analyses now allow connectivity between brain areas to differ over short periods of time. Consequently, connectivity changes can be assessed in the range of seconds instead of several minutes. By summarizing reoccurring large-scale patterns of connectivity, dFNC analyses can then be presented as distinct *connectivity states* of the brain, as well as transition trajectories between them. These dynamic measures may enable a more sophisticated evaluation of the spontaneously fluctuating nature of neural signals compared to static ones, may possess behavioral relevance (Vidaurre *et al*., 2019) and are increasingly suggested as novel biomarkers of disease (for recent reviews, c.f.: Hutchison *et al*., 2013; Preti *et al*., 2017; Lurie *et al*., n.d.). For example, Kim and colleagues (2017) applied dFNC analyses on Parkinson’s disease patients’ data and substantiated a significant association between the occurrences of dynamic connectivity states and clinical disease severity. These dynamic patterns uncovered reductions in functional network segregation that did not manifest in static analyses. Likewise, Espinoza and colleagues (2019) reported a significantly increased occurrence of a particularly weakly connected dynamic connectivity state in case of Huntington’s disease. This state’s dynamic connectivity diverged markedly from the well-known static one, thus facilitating previously inaccessible insights.

Given the increased capacity of dFNC analyses in delineating spontaneously forming connectivity states, such a dynamic approach seems to be particularly well suited to assess conditions requiring high levels of network flexibility, as, for example, in case of stroke-induced acute lesions. A focal lesion may not only disrupt communication within the motor system, but also alter the brain’s predilection for certain connectivity states (Vergara *et al*., 2018). Specific dynamic patterns may be crucial for the process of neural reorganization and hence determine the potential of the brain to recover (van der Horn *et al*., 2019).

Therefore, the goal of the current study was to investigate dynamic functional network connectivity of the motor system in acute ischemic stroke patients. We analyzed resting-state fMRI data from 31 first-ever stroke patients, presenting with moderate to severe hand motor deficits, and 17 age-matched healthy controls. We hypothesized distinct dFNC patterns in stroke patients linked to symptom load, i.e., specific patterns for severely and moderately affected patients, that were not assessable in the static analysis framework. In particular, we expected to observe temporally highly variable global changes in functional connectivity, readily interpretable as segregation and integration between functional domains (Friston, 2002; Eickhoff and Grefkes, 2011). These findings could then extend our mechanistic insights beyond the consistently described decrease in inter-hemispheric connectivity and thereby reveal new insights in the functional role of dynamic interactions among brain areas during the recovery process.

## Materials and Methods

### Participants

Thirty-two first-ever acute ischemic stroke patients admitted to the University Hospital of Cologne, Department of Neurology (mean age: 68.4 years±12.1 standard deviation (SD), 19 males, days post stroke: 7.2±2.7SD) and 17 healthy controls (mean age: 65.4 years±6.4SD, 15 males) were recruited for this study. One stroke patient had to be excluded due to severe head motion during scanning (see below), leaving 31 patients for the final analysis. Stroke patients presented with acute unilateral hand motor deficits. Further inclusion criteria were: i) 40–90 years of age; ii) diffusion-weighted magnetic resonance imaging (DWI) positive for ischemic stroke; iii) structurally intact ipsilesional pre-central gyrus (M1) as verified by MRI; iv) within two weeks from symptom onset (one patient was included 16 days after stroke); v) absence of severe aphasia, apraxia, and neglect. Exclusion criteria were: i) any contraindication to MRI (e.g., cardiac pace-maker); ii) epilepsy; iii) infarcts in multiple territories; iv) hemorrhagic stroke and v) further neurological diseases. In addition, 17 age-matched healthy subjects with no neurological or psychiatric disease served as control group. All subjects provided informed written consent in accordance with the Declaration of Helsinki and all aspects of this study were approved by the local ethics committee.

Please note that the raw data of 26 patients was previously included in Volz *et al*. (2016). Importantly, the scopes of the two studies strongly differ (dynamic functional connectivity analysis in acute stroke in the current study versus effects of repetitive transcranial magnetic stimulation (rTMS) on motor recovery in Volz *et al*. (2016)). There is no overlap, neither in the research question nor in any of the results. Therefore, all analyses described here are novel.

### Hand motor function and clinical assessments

Hand motor deficits were quantified using the Action Research Arm Test (ARAT) (Yozbatiran *et al*., 2008). This test is widely used in stroke research and assesses gross and fine upper limb function in four dimensions (i.e., grasp, grip, pinch, and gross movements; range 0–57; 57=normal performance, 0=unable to perform any movements). Furthermore, we obtained the National Institutes of Health Stroke Scale (NIHSS) for each patient.

In order to test for the impact of the motor deficit on dFNC, we divided the sample into i) a subgroup of severely affected patients (ARAT Score 0–28) and ii) a subgroup of lightly to moderately affected patients (ARAT score 29–57). For two patients, ARAT scores were unavailable due to technical reasons. The first patient was severely affected, according to an NIHSS of 16, and suffered from hemiplegia with no residual arm function. This was equivalent to an ARAT score close to zero, hence the patient was assigned to the severely affected subgroup.

The second patient was mildly affected (NIHSS=3) and had only minor hand motor deficits. Therefore, the patient was assigned to the moderately affected subgroup. Demographic characteristics of all study participants and clinical features of stroke patients are listed in **Table 1**.

**Table 1.** Demographics and clinical characteristics of stroke patients and healthy controls. Age and mean framewise displacement values were compared by means of two-sided *t*-tests, sex frequencies by means of a Pearson’s chi-squared test.

|  | **Stroke patients**<br>**(n=31)** | **Healthy controls**<br>**(n=17)** | **p-value** |
| --- | --- | --- | --- |
| **Age (in years)** | 68.4 | 65.4 | 0.4 |
| **Sex (in % female)** | 39 | 12 | 0.1 |
| **Mean Framewise Displacement** | 0.28 | 0.27 | 0.8 |
| **Days since stroke** | 7.2 | - | - |
| **NIHSS** | 8 | - | - |
| **ARAT *affected hand*** | 30 | - | - |
| **Lesion volume (in mm<sup>3</sup>, median)** | 12816 | - | - |
| **CST overlap (in %, median)** | 6.8 | - | - |

### Magnetic resonance imaging

Acquisition of resting-state functional MRI data was performed on a Siemens Trio 3T scanner (Siemens Medical Solutions, Erlangen, Germany). The following gradient echo-planar imaging (EPI) parameters were applied: repetition time (TR) = 2200 ms, echo time (TE) = 30 ms, field of view (FOV) = 200 mm, 33 slices, voxel size: 3.1 × 3.1 × 3.1 mm3, 20% distance factor, flip angle = 90°, 183 volumes (slice coverage of the whole brain). Participants were instructed to lie motionless in the scanner and keep fixating a red cross on a black screen all through the approximately seven minute session. For stroke patients, we additionally collected diffusion-weighted MR-images (DWI, TR = 5100 ms, TE = 104 ms, FOV = 230 mm, 30 slices, voxel size = 1.8 × 1.8 × 3.0 mm3) to obtain detailed information on lesion location and extent.

### Preprocessing of resting-state functional MRI data

Resting-state functional MRI data was preprocessed using the Statistical Parametric Mapping software package (SPM12, Wellcome Trust Centre for Neuroimaging, London, UK; www.fil.ion.ucl.ac.uk/spm) as implemented in MATLAB (version R2019a, MathWorks, Inc., Natick, MA, USA). In case of right-hemispheric lesions (n=8), images were flipped along the midsagittal plane. In this way, all lesions were located in the left hemisphere, thereby ignoring effects specific to the left or right hemisphere. The first four volumes (“dummy” images) of each scan were discarded to allow for the development of a steady blood-oxygenation level dependent (BOLD) activity signal. Preprocessing of the remaining 179 volumes continued with head movement correction by affine realignment to each scan’s mean image. In stroke subjects, diffusion-weighted images were co-registered to the mean EPI. Image volumes were spatially normalized using the “unified segmentation” option after masking lesioned tissue (Ashburner and Friston, 2005). In a final preprocessing step, images were smoothed using a Gaussian kernel with a full-width at half maximum (FWHM) of 8 mm.

### Controlling for head motion

We verified the absence of severe motion by calculating individual mean and maximum framewise displacements (Power *et al*., 2012). As dynamic functional connectivity analyses are sensitive to head motion, we excluded one subject (subject 32) with a maximum framewise translation of 12.2 mm. No significant group difference in framewise displacements was observable when comparing the final sample of 31 stroke patients with the 17 healthy controls (two-sided *t*-test: *p*=0.79).

### Intrinsic connectivity networks

The intrinsic connectivity network components used for dynamic functional connectivity estimations were computed using spatially constrained independent component analysis (ICA) (Lin *et al*., 2010; Du and Fan, 2013) based on components estimated from resting-state fMRI data of 405 healthy controls (Allen *et al*., 2014; Calhoun *et al*., 2001, components are available for download here: http://trendscenter.org/software/). The advantage of using a spatially constrained ICA approach with components from healthy participants on the current data sample is that parcellations are not biased by stroke lesions, yet are still adaptive to the individual and provide enhanced robustness to artifacts and noise compared to single-subject ICA denoising and regression based back-reconstruction (Salman *et al*., 2019; Du *et al*., 2016). We refer to Salman *et al*. (2019) for a detailed description of the group information guided ICA (“back-reconstruction”) algorithm. Given that all patients were scanned very early after stroke (on average 7.2 days), cortical reorganization is rather unlikely to have significantly changed the functional architecture (Rehme *et al*., 2011), further justifying the use of a healthy sample for defining the network components of interests.

As all patients were selected based on the presence of a motor deficit, we focused our analysis on the motor system. Accordingly, we obtained 13 motor network components (as extracted in Allen *et al*., 2014). These network components were identified as i) left (ipsilesional) primary sensorimotor cortex, ii) right (contralesional) primary sensorimotor cortex, iii) bilateral ventral premotor cortex, iv) supplementary motor area (SMA), v) bilateral postcentral gyrus, vi-vii) paracentral gyrus I and II and viii) bilateral superior parietal lobule; three subcortical (SC) components ix-xii): Putamen I, Putamen II and thalamus; and finally xii-xiii) two cerebellar (CB) components: cerebellum right and left. These network components were assigned to one of three domains, i.e., sensorimotor, subcortical and cerebellar domains (**Figure 1**).

**Figure 1.**
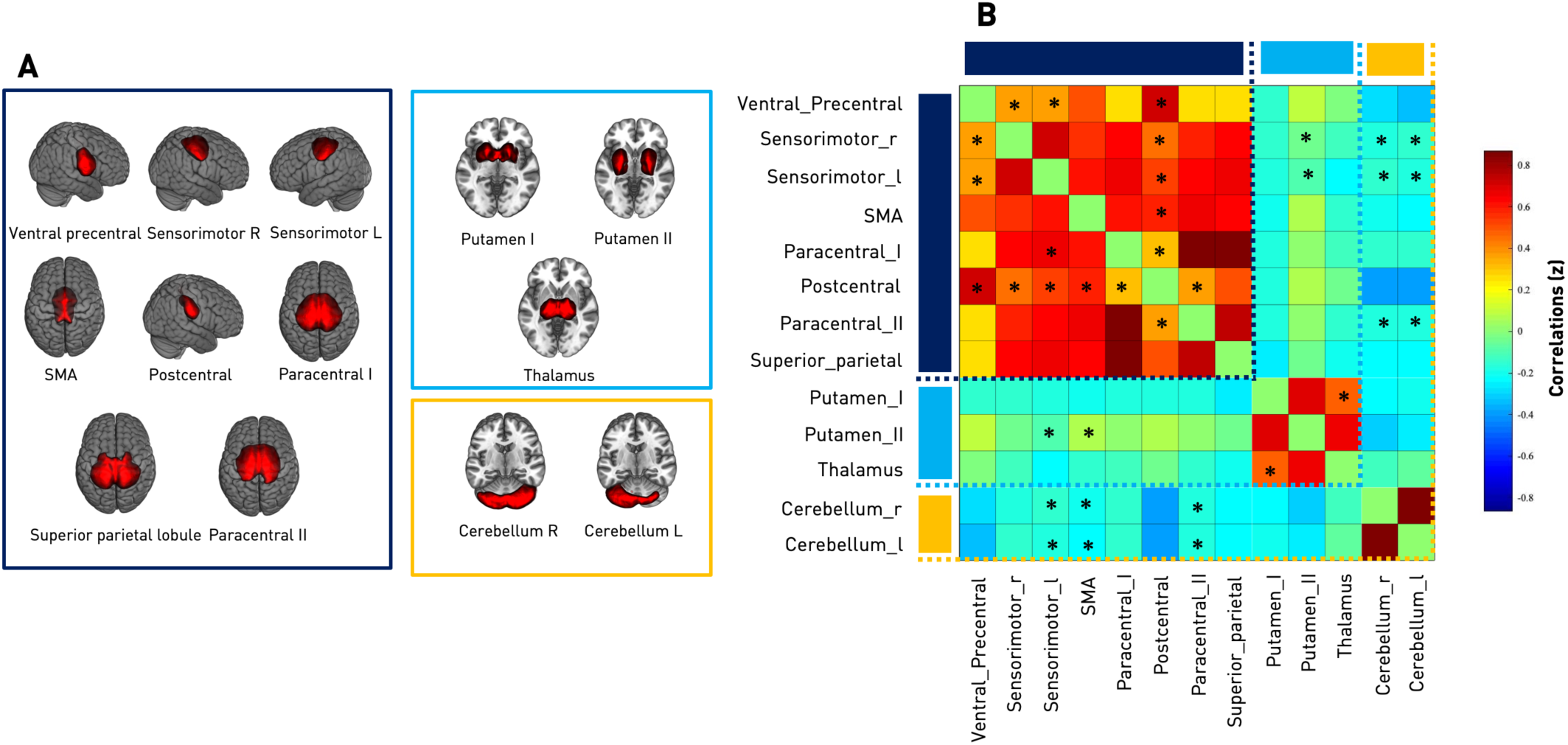
**A Spatial maps of 13 included independent components of all subjects (31 ischemic stroke patients and 17 healthy controls).** These were organized in three domains: Sensorimotor (SMN, 8 components, framed in dark blue), subcortical (SC, 3 components, framed in light blue) and cerebellar (CB, 2 components, framed in orange). Components were back-reconstructed based on the independent components of the cortical and subcortical sensorimotor as well as cerebellar domains presented in Allen *et al*., 2014. **B *Static* functional network connectivity between independent components resulting in a total of 78 connectivity pairs**. Connectivity values correspond to the Fisher’s Z-transformed Pearson correlation, averaged over the entire group of healthy controls and ischemic stroke patients. Red color indicates positive, blue color negative correlation. Thus, the connectivity matrix illustrates high positive intra-domain connectivity within the SMN, SC and SB domains as well as neutral to negative inter-domain connectivity: SMN-SC, SMN-CB and SC-CB. Asterisks indicate significantly altered static connectivity between the three subgroups: healthy controls, moderately and severely affected stroke patients (one-way ANOVA, *p*<0.05). The components “left ipsilesional sensorimotor” and “bilateral postcentral gyri” were both characterized by the highest number of disturbed *static* connectivity pairs (six each).

Before entering the resulting spatial maps and corresponding time-courses into further static and dynamic functional connectivity pipelines, time courses were detrended (i.e., accounting for linear, quadratic and cubic trends in the data), despiked using 3Ddespike (Cox, 1996) and filtered by a fifth-order Butterworth low-pass filter with a high-frequency cutoff of 0.15 Hz. Finally, each time-course was normalized for variance (Rachakonda *et al*., 2007).

### Static functional network connectivity analysis

The multivariate analysis of covariance (MANCOVAN) toolbox within GIFT (http://trendscenter.org/software/gift) was used to evaluate significant associations of connectivity within and between functional networks and the variables age, gender, translation, rotation and patient status (healthy controls – stroke patients). We proceeded in sequential multi- and univariate manners, as first outlined in Allen *et al*. (2011). Translation and rotation were computed as the mean of the absolute differences between consecutive time frames. Age, translation and rotation were treated as continuous variables, while gender and patient status were defined as discrete variables.

#### Within-network connectivity

Analyses were performed separately for each of the 13 network components’ spatial maps. We started with the full set of independent variables and PCA-dimensionality reduced voxel intensities of spatial maps as multivariate, dependent variables. We then obtained reduced models by backward elimination, i.e., relying on *F*-tests at each step, and retained significant independent variables only. Afterwards, we computed univariate *t*-tests for significant independent variables and thus probed the intensity difference for each voxel within a spatial map depending on a specific variable, while fixing remaining variables. As this resulted in several thousand individual *t*-tests, we applied false discovery rate (FDR)-correction for multiple comparisons to determine statistical significance with a level of *p*<0.05 (Benjamini and Hochberg, 1995).

#### Between-network connectivity

For each participant, we calculated static functional network connectivity strengths between components via Fisher’s Z-transformed Pearson’s pairwise correlation of time-courses. This resulted in 78 connectivity pair values per participant, according to the formula: 13 network components × (13 network components – 1) / 2 = 78.

We refined univariate tests from the two (healthy – stroke patients) to a three groups comparison (healthy – moderately affected – severely affected stroke patients) and thus evaluated static functional network connectivity differences in a three-level one-way ANOVA (level of significance *p*<0.05). Post-hoc *t*-tests (healthy – moderate, healthy – severe, moderate – severe) were performed in case of significant ANOVA results (level of significance *p*<0.05, FDR-corrected).

### Dynamic functional network connectivity

Dynamic FNC was estimated by means of the sliding window approach as implemented in the GIFT toolbox (Allen *et al*., 2014; Damaraju *et al*., 2014; Sakoğlu *et al*., 2010; Calhoun *et al*., 2014): We first defined 159 individual tapered windows by sliding time rectangles of 44 seconds (width=20 TRs). These time windows were convolved with a Gaussian of seven seconds (σ=3 TRs) and shifted in steps of 2.2 seconds (1 TR). We opted for common parameter settings, particularly as prior studies have provided evidence that a window width between 30 to 60 seconds enables successful derivation of dFNCs, which are not governed by noise (Liégeois *et al*., 2016; Preti *et al*., 2017). Within each of these windows, we computed dFNCs from the *l*1-regularized precision matrix, i.e. sparse inverse covariance matrix (Friedman *et al*., 2008; Varoquaux *et al*., 2010; Smith *et al*., 2011). We regressed out the covariates age, sex, mean framewise translation and rotation. Finally, we applied Fisher’s Z-transformation to functional connectivity matrices to obtain Z values and stabilize variance for further analyses.

### Clustering analysis

Aiming to condense and thereby improve the interpretability of the rich information of 159 dFNC matrices per subject, we used k-means clustering (Lloyd, 1982) to compute re-occurring functional connectivity patterns, i.e. *connectivity states*, across time and subject space (Hutchison *et al*., 2013; Allen *et al*., 2014; Calhoun *et al*., 2014). We implemented the *l*1 distance function (“Manhattan distance”) given its suitability for high-dimensional data (Aggarwal *et al*., 2001). As is convention (Allen *et al*., 2014; Espinoza *et al*., 2018), this clustering approach was applied to all subjects’ connectivity matrices twice: Firstly, to determine the optimal number of clusters *k (*referred to as states), and then to construct the final *k* connectivity states. Each window of each subject was assigned to one of these connectivity states. Notably, this analysis does not guarantee that all participants visit all of the connectivity states, i.e. some might enter only one or two states in spite of three or more existing states. We compared the group-averaged connectivity states originating from the dynamic functional connectivity analysis with the group-averaged static functional connectivity obtained earlier by calculating the Manhattan distance.

### Statistics of dynamic connectivity measures

We statistically evaluated dynamic connectivity specific measures: i) *Fraction times* (the percentage of total time a subject spent in a state), ii) *dwell times* (the time a subject spent in a state at a time without switching to another one in the meantime), iii) *numbers of transitions* (how often a subject changed states) and iv) *transition likelihoods* (percentages of transition likelihood between the *k* connectivity states). Additionally, we tested for group differences in dynamic connectivity pairs within each connectivity state. Similar to the between-network static functional connectivity analysis, we initially performed three-level one-way ANOVAs to test for differences between the three groups: healthy controls, moderately and severely affected stroke patients (level of significance *p*<0.05). Post-hoc *t*-tests (healthy – moderate, healthy – severe, moderate – severe) were added in case of significant ANOVA results (post-hoc *t*-tests: level of significance, *p*<0.05, FDR-corrected).

### Data and code availability

Matlab scripts for dFNC computation were based on templates available in the GIFT toolbox, additional jupyter notebooks in python 3.7 for statistical evaluations and visualizations can be found here: https://github.com/AnnaBonkhoff/DFNC_Stroke.

## Results

### Demographic and clinical characteristics

There were no significant differences in age, sex category and mean framewise displacement between healthy controls and all stroke patients (**Table 1**). Patients were scanned 7.2 (±0.6 Standard Error of the Mean (SEM)) days after stroke onset with no statistically significant difference in time since stroke between subgroups.

Since we defined patient subgroups based on a discrete ARAT-cut-off of 29 (i.e., half of the maximum score), severely affected patients (n=13, ARAT<29) and moderately affected patients (n=18, ARAT≥29) differed significantly in regard to their motor performance (*p*<0.05): Arm motor function was significantly lower in the severely affected group (ARAT=10.0±3.1SEM) compared to the moderately affected group (ARAT=44.5±2.4SEM). In contrast, covariates including age, days post-stroke and lesion volume did not significantly differ between groups (**Supplementary Table 1**).

### Static functional network connectivity

#### Within-network connectivity

MANCOVAN analyses indicated statistically significant group differences between all stroke patients and healthy controls within the spatial maps of all eight cortical sensorimotor components, two subcortical components (Putamen I & II, yet not the thalamus) and one cerebellar component (right cerebellum). Subsequent univariate analyses, centering on the variable patient status (healthy controls – stroke patients) and signal intensities of each voxel within a spatial map, provided further evidence of significantly lowered signals within the outlined components in case of stroke patients (**Figure 2**).

**Figure 2.**
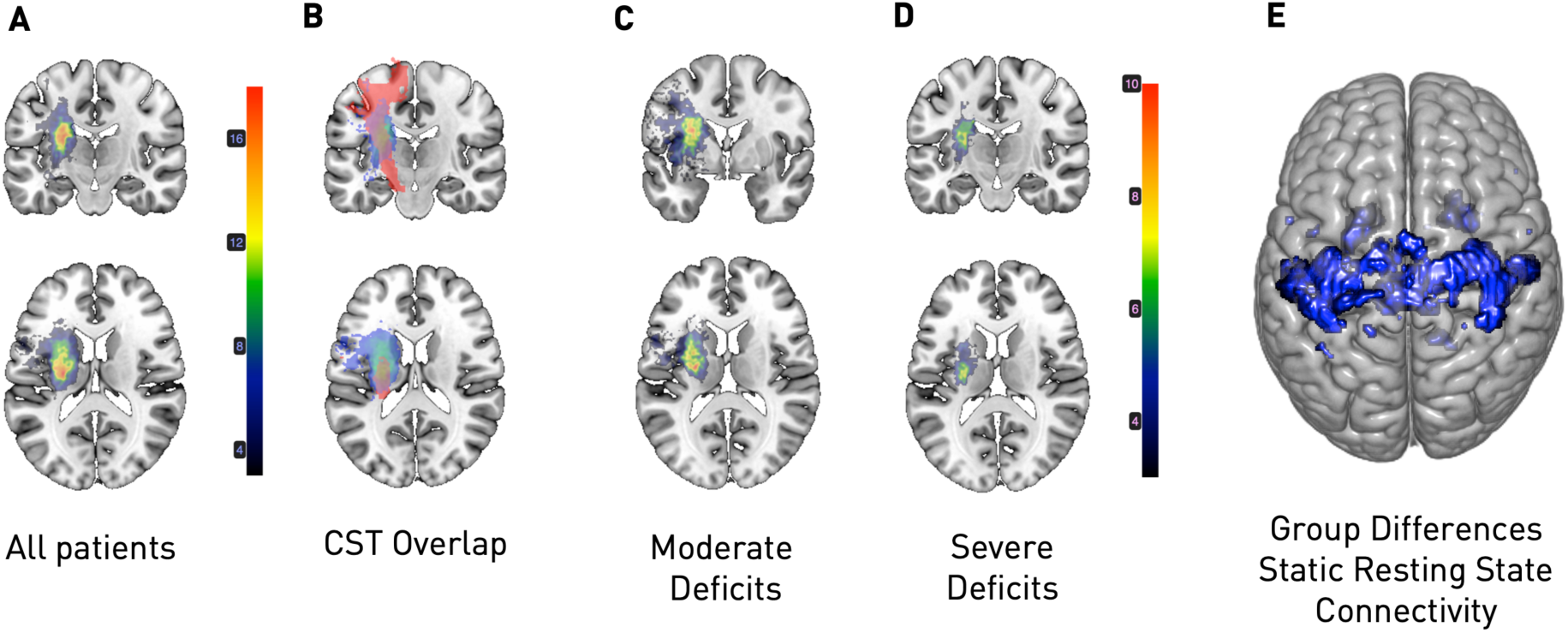
Overlap maps of DWI lesions (A-D) and within-network connectivity (E). **A** Entirety of stroke patients: The majority of lesions was located subcortically, with the maximum overlap (n=18/31) being in the posterior limb of the internal capsule. **B** Illustration of the lesion overlap to the corticospinal tract (CST, in red, median overlap for the entire stroke patient sample: 6.8%). **C** Subgroup of moderately affected stroke patients (n=18). **D** Subgroup of severely affected stroke patients (n=13). Both stroke patient subgroups primarily presented with subcortical lesions. Importantly, while subgroups were defined based on their motor function (cut-off: ARAT 28/29), they neither differed in lesion volume, nor corticospinal tract overlap, c.f. **Supplementary table 1. E** Stroke patients exhibited significantly reduced within-network connectivity in multiple sensorimotor cortical and subcortical components compared to healthy controls, i.e., bilaterally in the precentral gyri, the postcentral gyri, the supplementary motor area, paracentral gyri, superior parietal lobule and subcortically in the Putamen (*p*<0.05, FDR-corrected for multiple comparisons). This finding, therefore, replicates main observations in previous studies on functional connectivity alterations post-stroke (e.g., Carter et al., 2010, Golestani et al., 2013, Rehme et al., 2014).

#### Between-network connectivity

Static functional network connectivity (over the entire scanning time series) was calculated as (Fisher Z-scored) Pearson’s correlation between 78 individual component pairs: In general, we observed strong intra-domain connectivity, i.e. component pairs within either the sensorimotor, subcortical or cerebellar domains were highly positively correlated. In contrast, inter-domain connectivity was comparably low, i.e., components from sensorimotor and subcortical or cerebellar domains were either independent from each other or negatively connected (**Figure 1(B)**). When screening for group differences between controls, moderately and severely affected patients by means of a one-way ANOVA, 18 connectivity pairs showed significantly altered between-network component connectivity. These alterations were mostly located in the cortical sensorimotor domain (**Figure 1(B)**). Post-hoc *t*-tests, contrasting severely affected patients and healthy controls, revealed a stroke-induced increase in connectivity between subcortical components (*p*<0.05, FDR-corrected). In contrast, moderately affected patients comprised decreases in connectivity strength when compared to healthy controls (*p*<0.05, FDR-corrected). For example, connectivity was lower between the left and right components and ventral precentral components, as well as postcentral components and the supplementary motor area and both cerebellar components (c.f., **Figure 3** for details on altered connectivity pairs). Moderately and severely affected patients did not feature significantly different static connectivity after correction for multiple comparisons.

**Figure 3.**
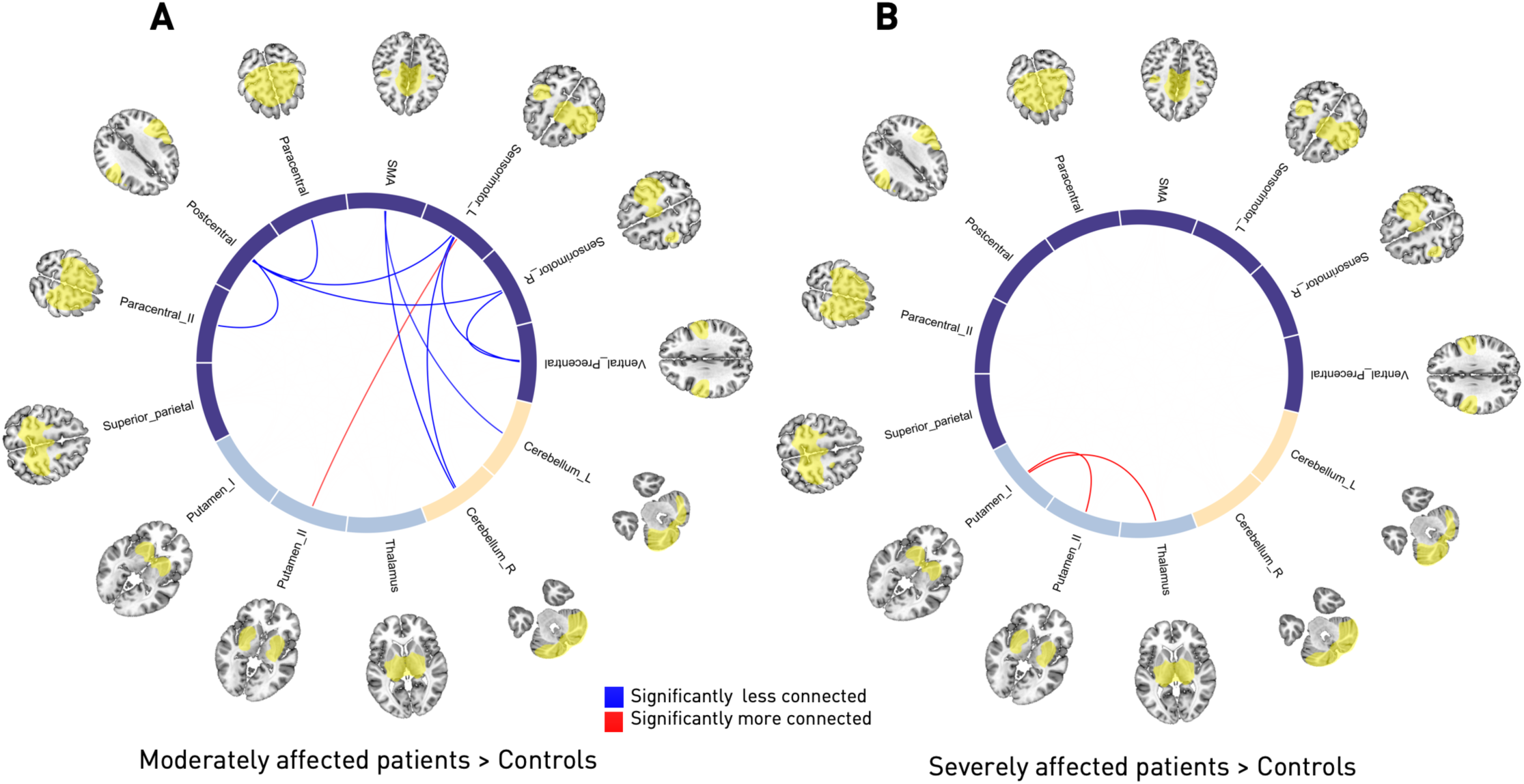
Circle plots of significant static functional connectivity differences between the subgroups (post-hoc *t*-tests, *p*<0.05, FDR-corrected for multiple comparisons). **A Healthy controls versus moderately affected stroke patients.** Connectivity strength in stroke patients was found to be decreased between the pre- and postcentral areas and between the supplementary motor area and bilateral cerebellar components as well as the left, ipsilesional precentral gyrus and the right cerebellum, yet comparably increased between the ipsilesional precentral gyrus and Putamen. **B Healthy controls versus severely affected stroke patients**. In contrast to the previous group comparison, only two significantly altered connectivity pairs emerged: The connectivity between both of the Putamen components as well as the more anterior Putamen component with the thalamic component were increased in stroke patients. Note that there were no significant connectivity differences between moderately and severely affected stroke patients.

### Dynamic functional network connectivity

We next investigated the temporal properties of functional connectivity, i.e., dFNC. By applying the k-means clustering algorithm to the estimated 159 functional connectivity matrices per subject and the optimization criteria for the number of states mentioned above, we identified three connectivity states, i.e., quasi-stable connectivity patterns, that re-occurred across all subjects during scans collection (**Figure 4(B)**). The states are presented and described in the order given by k-means. The first connectivity state was characterized by highly positive intra-domain connectivity and highly negative inter-domain connectivity. We refer to this state as the *regionally densely connected* state with strong inter-domain segregation (overall frequency of State 1: 29%, **Figure 4(A)**). This connectivity state was also the one that most closely matched the static connectivity estimates in terms of Manhattan distance. The second connectivity state featured comparably weak intra-domain connectivity, which was particularly true for connections of the ventral precentral and postcentral components. Inter-domain connectivity values close to zero additionally implied a low connectivity between the three domains. We therefore call this state the *weakly connected* state (overall frequency of State 2: 43%, **Figure 4(A)**). The third connectivity state represented a combination of the preceding two: Positive intra-domain connectivity, slightly positive connectivity between the sensorimotor and subcortical domains and negative connectivity between the cerebellar and both of the sensorimotor and subcortical domains (overall frequency of State 3: 27%, **Figure 4(A)**). Therefore, instead of *one* static connectivity state in the form of *one* FNC matrix, we now obtained *three* dynamic connectivity states and dFNC matrices. Importantly, one dynamic connectivity state, the *weakly connected* State 2, differed markedly from the static version.

**Figure 4.**
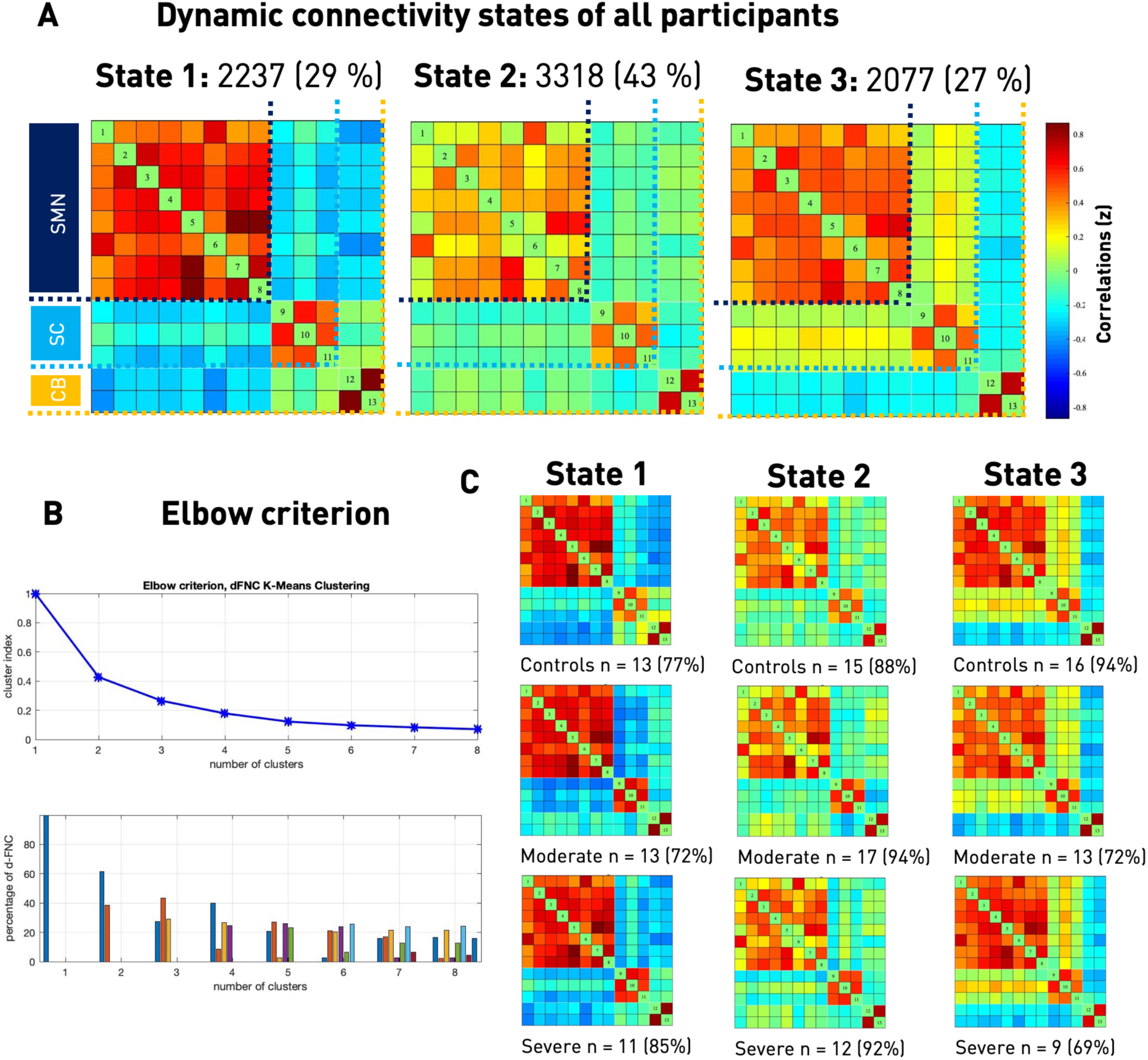
Dynamic functional network connectivity analysis. **A Three resulting connectivity states as well as their across-group frequencies.** The first state was characterized by a highly positive intra-domain connectivity in all of the domains (SMN, SC, CB) and highly negative inter-domain connectivity. It was the state resembling the static connectivity matrix the most, measured in Manhattan distance. The second and most frequent state featured comparably lower connectivity within the sensorimotor domain, particularly between the ventral precentral component and the paracentral lobule to further sensorimotor components. Inter-domain connectivity was mostly neutral. The third state comprised positive intra-domain connectivity, negative inter-domain connectivity between both the sensorimotor and subcortical domains to the cerebellar domain and no connectivity between the sensorimotor and subcortical domains. **B Elbow criterion**. Trajectory of the clustering index with increasing numbers of clusters, i.e., *k* in k-means clustering (upper row) and cluster distributions for a given number of clusters (bottom row). As the steepness of the decline in clustering index decreased markedly after three and four clusters, yet the four cluster solution included a state with a frequency of <10%, k=3 combined the lowest clustering index and most well-balanced solution. **C Connectivity states separately for each of the three subgroups**. Please note that some subjects only entered one or two of the defined three connectivity states, resulting in varying numbers of subjects within a specific state (c.f., stated absolute numbers of subjects entering the state as well as the percentage of the entire subgroup). Connectivity frequencies did not differ significantly between subgroups and states.

### Temporal characteristics

We subsequently tested for between-group differences in the measures of dynamic behavior. Three-level one-way ANOVAs comparing healthy controls, moderately and severely affected stroke patients revealed significant differences of temporal features in State 2, i.e., the *weakly connected* state (Fraction and Dwell times of State 2: *p*<0.05, **Figure 5(A), (B)**). Moderately affected patients had the most deviating behavior: When contrasted with healthy controls, they particularly preferred State 2 and spent more time in it in total as well as dwelled longer once having entered it (post-hoc t-tests: Fraction and Dwell times *p*<0.05, FDR-corrected). With respect to severely affected patients, moderately affected patients again tended to spend more time in State 2 (post-hoc *t*-tests: Fraction time: *p*=0.07, FDR-corrected; Dwell time: not significant). In contrast, there were no significant between-group differences for the absolute number of transitions between states. Subjects switched five to seven times on average during the entire scanning period (one-way ANOVA: *p*>0.05, **Figure 5(C)**).

**Figure 5.**
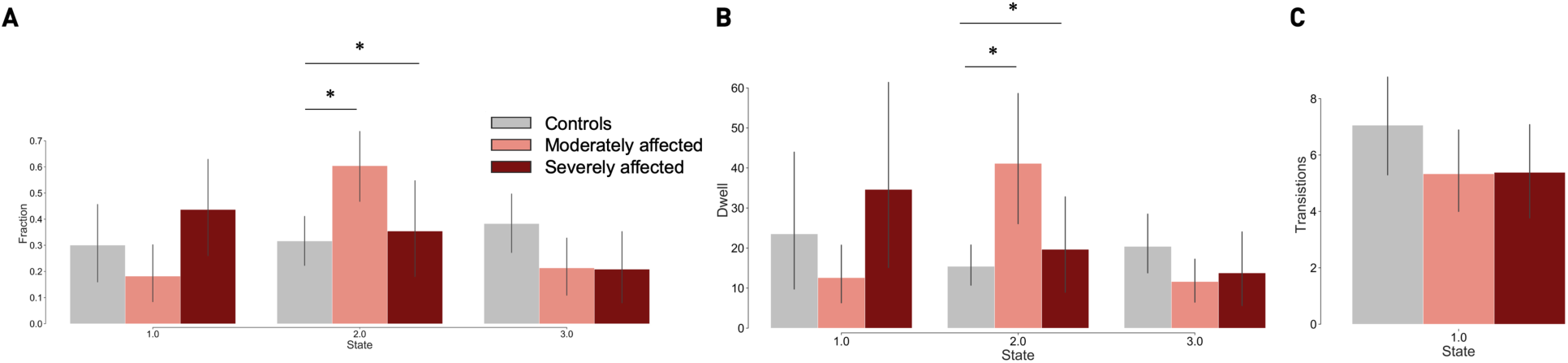
Dynamic functional network connectivity analysis: Fraction and dwell times as well as the number of transitions for the three groups: healthy controls, moderately and severely affected patients (asterisks indicate statistically significant group differences based on one-way ANOVAs, and post-hoc *t*-tests, *p*<0.05, FDR-corrected for multiple comparisons). **A Fraction times**. Over the entire scan duration, moderately affected stroke patients spent significantly more time in State 2 than healthy controls. State 2, the generally most frequent connectivity state, was characterized by comparable low positive intra-sensorimotor domain connectivity. **B Dwell times**. Once again, moderately affected stroke subjects differed from healthy controls and spent significantly more time in State 2 at any one time. **C Number of transitions**. The absolute number of transitions did not differ significantly between the three groups. Subjects switched states between five to seven times on average.

Next, we evaluated the transition likelihoods between states. There were significant between-group effects (ANOVA *p*<0.05) with respect to the likelihood of staying within a state and switching to a new one: Consistent with the observed increase in fraction and dwell times in State 2, moderately affected patients were more likely to stay in the *weakly connected* state compared to both healthy controls and severely affected patients (post-hoc *t*-tests: *p*<0.05, FDR-corrected). Of note, severely affected patients presented with a markedly different behavior: They were not only less likely to remain in the *weakly connected* State 2, but demonstrated a significant preference for the *regionally densely connected* State 1. Coming from State 2, they were more likely to switch to State 1 compared to moderately affected patients (post-hoc *t*-tests: *p*<0.05, FDR-corrected) and healthy controls (*p*=0.056, FDR-corrected, **Figure 6**). Moderately affected patients and controls did not differ in this aspect.

**Figure 6.**
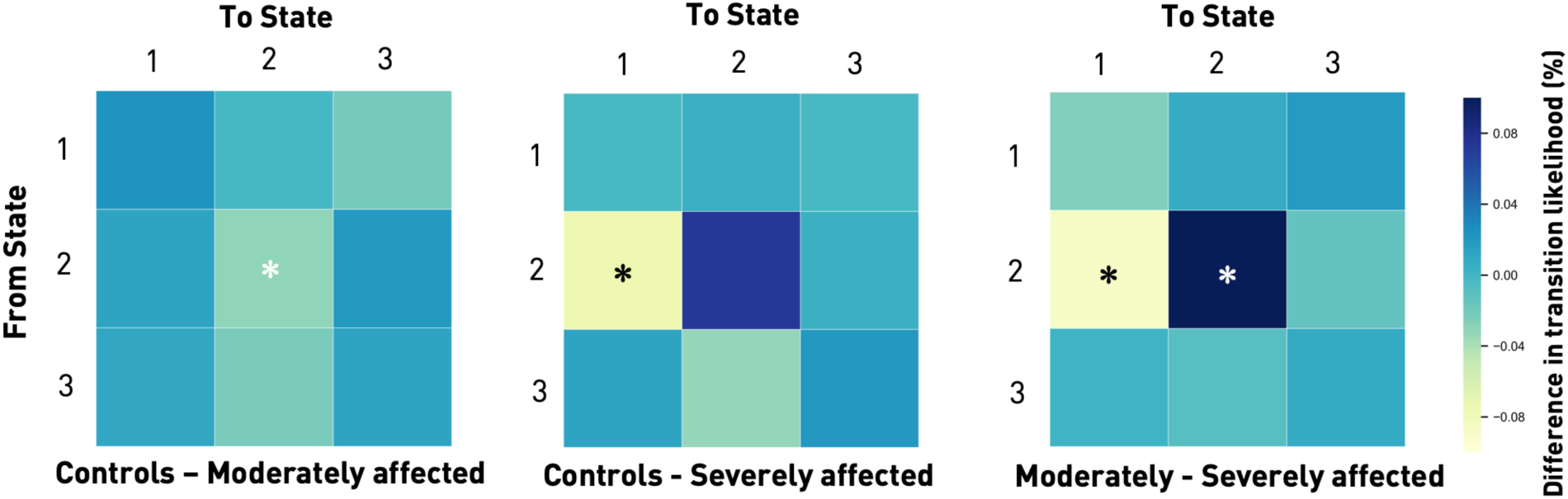
Transition matrices displaying the differences in likelihood of changing from one connectivity state to another between the subgroups. Generally, subjects tended to stay in their current connectivity state. Thus, if they were in State 1 at any one time *t*, they would most likely be in the same connectivity state at *t+1*, i.e., the next window. The same was true for States 2 and 3 (likelihood of remaining in the same state: range 87% - 97%). Transitions from any one state to one of the other two states were less likely (likelihood range 1 – 10%). There were statistically significant group differences for transitions from State 2 to itself as well as to State 1 (one-way ANOVAs, *p* < 0.05). Moderately affected stroke patients had a significantly higher likelihood of staying in State 2 than healthy controls and severely affected stroke patients (post-hoc *t*-tests, *p*<0.05, FDR-corrected for multiple comparisons). In contrast, severely affected stroke patients had a higher likelihood of not staying in State 2, but switching from this state to State 1 (post-hoc *t*-tests, *p*<0.05, FDR-corrected for multiple comparisons). Therefore, they preferably transitioned to the *regionally densely connected* State 1 and left the *weakly connected* State 2.

### Dynamic connectivity characteristics

Lastly, we examined between-group differences in connectivity strengths for each of the three connectivity states. While we did not detect any significant effects for State 1, i.e. the *regionally densely connected* state, moderately affected patients featured a number of differences in States 2 and 3 compared to both severely affected patients and healthy controls. This was especially true with respect to the connectivity of ipsilesional primary sensorimotor cortex and subcortical components (**Figure 7**). In moderately affected patients, connectivity differences in State 3 matched those of the static analysis to a great extent, with reduced connectivity in stroke patients between the ipsilesional left and contralesional right sensorimotor components, as well as bihemispherical SMA and one of the paracentral lobule components. Noteworthily, we found reduced connectivity between the right contralesional sensorimotor and paracentral lobule components when contrasting severely affected patients with controls. This inter-hemispheric difference was not detectable in the static analysis and can hence be interpreted as increased sensitivity of the dynamic analysis. Lastly, moderately and severely affected patients differed in three cortical–subcortical–cerebellar connectivity strengths: Paracentral – Putamen, left Sensorimotor – Putamen and Putamen – left Cerebellum (**Figure 7)**.

**Figure 7.**
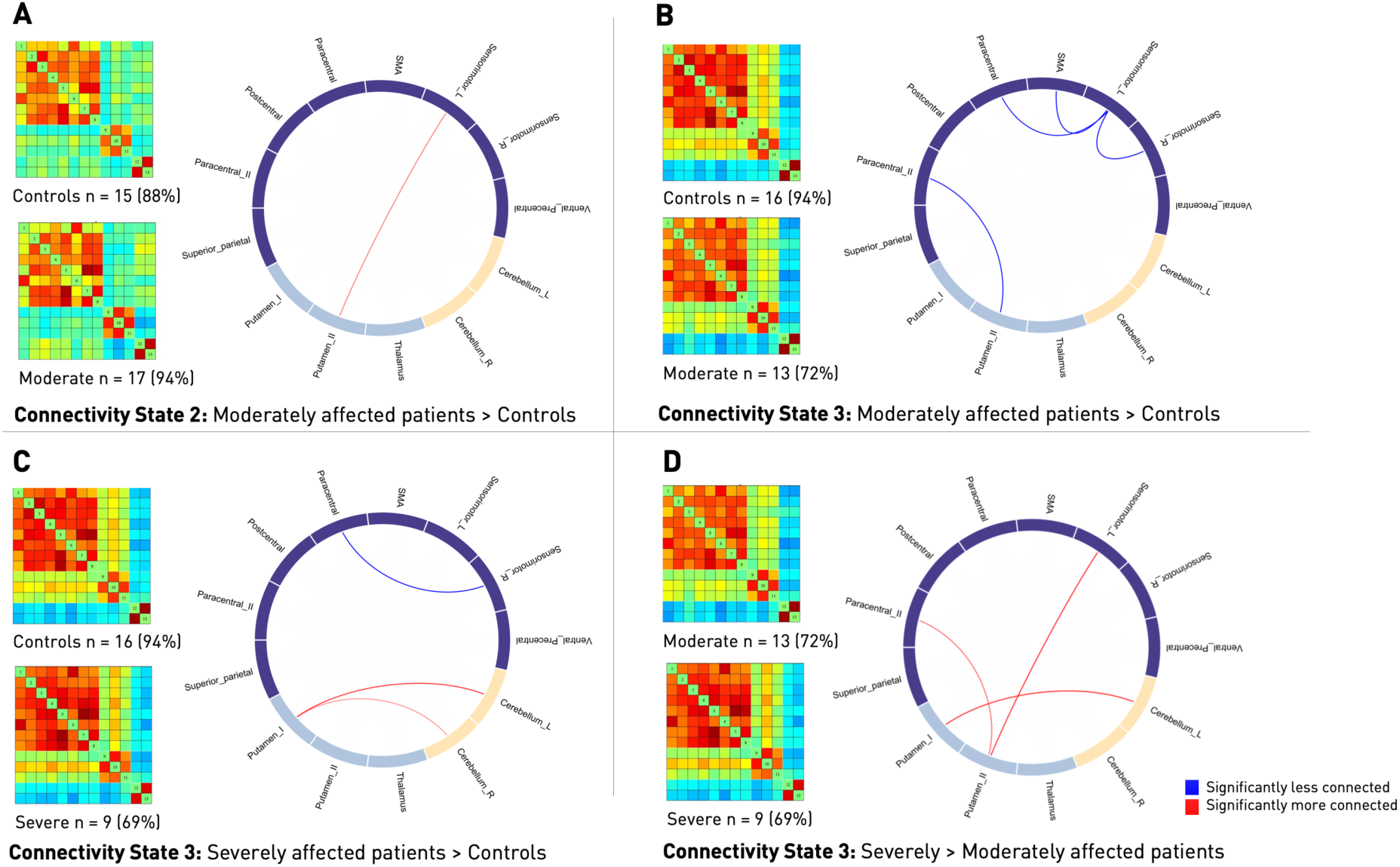
Differences in dynamic functional connectivity between the various subgroups: Subgroup-specific connectivity matrices as well as numbers and percentages of subjects within a group entering this state (left) and circle plots of significant functional dynamic connectivity differences (right, post-hoc t-tests, p<0.05, FDR-corrected for multiple comparisons). **A & B Differences between healthy controls and moderately affected stroke patients**. Moderately affected stroke patients presented higher connectivity strength between the ipsilesional sensorimotor area and the more posterior Putamen component, yet lower connectivity between a bilateral, more ventral precentral component and the same posterior Putamen component in State 2 (A). Furthermore, connectivity differences in State 3 resembled the ones in the static functional connectivity analysis: Stroke patients featured significantly lower connectivity between the ipsi- and contralesional sensorimotor areas as well as lower connectivity between the ipsilesional sensorimotor area and the paracentral lobule and supplementary motor area (B). **C Differences between healthy controls and severely affected patients in State 3**. Stroke patients presented with a decreased connectivity between the contralesional, right sensorimotor and paracentral areas, while connectivity between the more anterior Putamen component and both cerebellar components was increased. **D Differences between moderately and severely affected stroke patients in State 3**. Several connectivity pairs were reduced in case of moderately affected stroke patients. These pairs were: Ipsilesional, left sensorimotor cortex – Putamen, paracentral cortex – Putamen and the more anterior Putamen component – left cerebellum.

In summary, we found significant differences in preferences for connectivity states depending on the degree of motor impairment: While moderately affected patients spent more time in a *weakly connected* state, severely affected patients transitioned more often to a *regionally densely connected* state with strong intra-domain (e.g. cortico-cortical) connectivity and negative inter-domain (e.g. cortico-subcortical) connectivity. Interestingly, and somewhat surprising, functional connectivity estimates differed more between moderately affected patients and healthy controls than between severely affected patients and healthy controls. This was particularly the case for disturbed static functional connectivity between specific brain regions, where moderately affected patients presented with multiple reduced functional connectivity pairs. In case of severely affected patients and the static analysis, we found altered functional connectivity pairs only in the subcortical domain. Disturbances at the level of cortical regions only became evident in the dynamic analysis. We here substantiated lowered connectivity between the contralesional sensorimotor and paracentral lobule components post-stroke, indicating an added benefit of dynamic analysis.

## Discussion

Dynamic functional network connectivity analyses encompass the unique capacity of more fine-grained conclusions in the temporal domain (Allen *et al*., 2014; Calhoun *et al*., 2014). In this study, we dissected the dynamic connectivity behavior of ischemic stroke patients within the first few days of their acute event, putting a particular emphasis on hand motor function and the sensorimotor system. Of note, stroke patients diverged from healthy controls depending on the severity of their motor deficit: Patients with severe motor symptoms showed a significant preference for transitioning to State 1, a *regionally densely connected* state with strong intra-domain connectivity. Conversely, moderately impaired patients spent significantly more time in State 2, a *weakly connected* state. Both of these patterns remained hidden in previous conventional static analyses. The third connectivity state did not differ in terms of dynamic connectivity specific measures, such as transitions, fraction and dwell times, but reflected many of the connectivity differences observed in the static analysis and previously described in the literature (Carter *et al*., 2010; Wang *et al*., 2010; Golestani *et al*., 2013; Rehme *et al*., 2014). Contrary to our initial expectations, moderately affected patients comprised more pronounced alterations in functional connectivity for both the static and dynamic analyses, despite their weaker clinical deficit. These alterations were located between the ipsi- and contralesional sensorimotor cortices as well as the contralesional sensorimotor cortex and bilateral paracentral cortex. Importantly, in case of severe motor symptoms, significantly reduced inter-hemispheric connectivity between cortical sensorimotor components was exclusively found in the dynamic analysis, highlighting its higher sensitivity in comparison to the static analysis.

### Functional integration and segregation

Dynamic connectivity states can be interpreted within the concepts of *functional integration* and *functional segregation* (Friston, 2011; Eickhoff and Grefkes, 2011). Functional segregation refers to the parcellation of the brain into regionally unique *modules* (i.e., areas), each of which may be assigned to a particular domain. These domains can either orchestrate information transmission from one to the other – referred to as integration – or process information in isolation – referred to as segregation; essentially balancing the two extremes to maintain healthy brain functions (Sporns, 2013). In the following, we will evaluate the computed three connectivity states within the framework of these concepts, establish links to recent literature and embed our findings.

### Severely affected patients & State 1

The *regionally densely connected* State 1 was characterized by highly positive intra-domain connectivity, i.e. within the sensorimotor, subcortical or cerebellar domains, and negative inter-domain connectivity, i.e. between sensorimotor – subcortical, sensorimotor – cerebellar and subcortical – cerebellar domains. Altogether, this can be interpreted as high segregation between domains. In view of the significantly increased transition likelihood to this State 1, severely affected patients thus preferred a state where information could easily travel from one component to another in the same domain. However, information exchange between components of distinct domains was hindered. This high intra-domain connectivity or integration was reminiscent of the over-activation and excessive recruitment of cortical motor areas, particularly of contra-lesional M1, bilateral premotor cortex and SMA, in task-based studies post-stroke (e.g., Ward *et al*., 2003; Rehme *et al*., 2011, for a review c.f., Rehme *et al*., 2012). Importantly, Rehme *et al*. (2011) uncovered these activity changes *exclusively* for severely affected patients, starting a few days after stroke, and suggested these to represent early signs of reorganization. Conceivably, the observed *dynamic* pattern here, even though expressing connectivity instead of activation and recruitment, could be a respective correlate – demonstrating the increased employment of functional connections within well-defined domains, aiming to recover lost motor performance. This specific pattern may not have been discovered and described in preceding *static* analyses, as it was solely expressed in the temporal markers of our *dynamic* analysis.

Segregation, as a measure of spatial functional specialization, can be quantified in multiple ways and is often linked to brain modularity. This graph theoretical measure compares densities of connections within and between domains, with higher values implying greater modularity and segregation of domains (e.g., c.f., Newman, 2004). Brain modularity was recently suggested as biomarker of intervention-related brain plasticity, since higher baseline values of modularity were shown to be predictive of later gains in cognitive function (Gallen and D’Esposito, 2019). This relationship could be established not only for healthy aging brains, but also, notably, structurally damaged brains of traumatic brain injury patients (Arnemann *et al*., 2015; Gallen *et al*., 2016; Baniqued *et al*., 2018). Similarly, when limiting analyses to the cortical motor and visual domains, Mattar *et al*. (2018) extracted a link between pronounced modularity and prospective motor skill learning. In these regards, severely affected patients’ predilection for the *segregated* State 1 could represent an attempt to facilitate brain plasticity and regain motor performance. Nonetheless, it has previously been hypothesized that modularity and segregation might follow an inverse U-shaped curve and values on either end of the scale could co-occur with maladaptive behavior (Duncan and Small, 2016). Therefore, not having any available long-term behavioral markers prohibits a definitive conclusion at this point and necessitates future research to investigate links between segregation and motor recovery further.

### Moderately affected patients & State 2

In contrast, moderately affected patients spent significantly more time in State 2 that comprised low intra-domain integration and relatively increased inter-domain connectivity – which is interpretable as reduced segregation. Decreased segregation has been found in healthy as well as pathological aging (Chan *et al*., 2014; Wig, 2017; Kim *et al*., 2017). In principle, this pattern of decreased segregation is also well in line with previous reports of reductions in modularity post-stroke, reflecting a decrease in segregation between domains (Gratton *et al*., 2012; Duncan and Small, 2016; Siegel *et al*., 2016; Siegel *et al*., 2018). Duncan and Small (2016) and Siegel *et al*. (2018) reported initially reduced and subsequently increasing segregation in parallel to functional recovery, suggesting reduced segregation to be a signature of impaired function. However, substantial differences in average scanning time after stroke (i.e., two weeks (Siegel), forty months (Duncan & Small) and one week in our study) and clinical deficit (i.e., predominantly cognitive symptoms (Siegel), aphasia (Duncan & Small) and motor symptoms here) impede any direct transfer of their conclusions to our findings. The interpretation of reduced segregation in moderately affected patients as a correlate of impaired function is additionally challenged by the fact that clinically more affected patients did not show similar alterations. Hence, in our setting, reduced segregation rather appeared as a signature of (early) reorganization, given that moderately and severely affected patients differed in motor performance by definition, but not in lesion volume or corticospinal tract overlap. Also, one could imagine that new connections could be more easily established starting from a weakly connected state, providing a basis for higher network flexibility. In contrast, the *regionally densely connected* State 1 with high intra-domain, yet low inter-domain connectivity might facilitate regional processing and hence reorganization in the case of a severe disruption of motor output.

By describing dynamic functional connectivity characteristics of subcortical ischemic vascular disease (SIVD) patients, Fu and colleagues’ (2019) study may be one of the currently most comparable studies to ours, given the scarcity of dynamic functional connectivity studies investigating stroke patients. Clinically, SIVD patients predominantly present with prolonged and not necessarily acute onset neurocognitive decline as well as chronic deep white matter hyperintensities and subcortical lacunar infarcts (Hachinski *et al*., 2006). The analysis of their dynamic functional connectivity revealed that fraction and dwell times were increased in a weakly connected state and reduced in a densely connected state – the former finding being much alike our main finding for moderately affected patients. Duncan and Small (2018), on the other hand, leveraged the sliding window approach to track dynamic network changes of twelve chronically aphasic stroke patients (on average 40 months post-stroke) during a six week intervention. Estimating ten different connectivity states in total, the increase in dwell time of one of these, a particularly weakly connected state, correlated with an increase in language performance. Altogether, the previous and present findings suggest a consistent link between dynamic measures and vascular disease, rendering it a promising future technique in the field of neurorehabilitation.

### Limitations and future directions

Although our acute stroke patient sample size of n=31 is rather small, it was apparently equipped with sufficient statistical power, especially given the replication of previous static functional connectivity findings. We were able to exploit detailed measures of motor function by relying on a competitive group size of severely affected patients (n=13) (c.f., six to ten severely affected patients in Carter *et al*., 2010; Wang *et al*., 2010; Park *et al*., 2011; Golestani *et al*., 2013; Rehme *et al*., 2014). We further link current limitations to future directions: In light of the spatial heterogeneity of stroke lesions as well as not yet satisfactorily accurate predictions of individual recovery trajectories, future studies should involve larger sample sizes and evaluate symptoms in a fine-grained as well as prospective fashion to corroborate and extend current finding. In this way, confidence in their generalizability could be increased. Eventually, this would represent a natural succession to our approach presented here, where we focused on dynamic connectivity differences within motor-related networks at the acute stage post-stroke. It would enable the elucidation of pressing questions, such as: Are dynamic connectivity measures predictive of future outcomes, especially in regard to treatment effects, including transcranial magnetic stimulation (TMS) and transcranial direct current stimulation (tDCS)? How do non-motor symptoms affect dynamic connectivity comprising all conceivable domains? How do dynamic connectivity measures evolve over time? Existing literature already suggests a delicate relationship between connectivity changes and the exact time point post-stroke: It has been shown that asymmetries in bi-hemispherical connectivity peaked approximately one month after the acute event (Park *et al*., 2011) and, considering rats, different temporal trajectories emerged depending on lesion location (van Meer *et al*., 2010). Asynchronistic behavior could conceivably explain why severely and moderately affected patients differed in their dynamic connectivity profiles in our analysis. As a reasonable interpretation, moderately affected patients may have traversed through a more segregated phase already when regaining function, while severely affected patients lag behind. Lastly, it might be promising to link findings of fMRI determined connectivity states to micro states as inferred from EEG-data, in light of the increased temporal resolution in both cases (with EEG still providing higher resolutions, c.f., Zappasodi *et al*., 2017).

## Conclusion

Dynamic functional connectivity analyses hold promise to capture critical characteristics of cognition and behavior in health and disease (Hutchison *et al*., 2013). In this study, we build upon this notion by implementing dynamic functional connectivity analyses to investigate the temporal behavior within the motor system of 31 acute stroke patients presenting with impairments of hand motor function. Significant deficit-severity dependent dynamic patterns demonstrated the added value of dynamic connectivity measures as (resting-state) biomarkers of ischemic vascular disease: Patients with severe deficits preferred a *regionally densely connected*, highly segregated state, conceivably representing an effort to induce brain plasticity and subsequent motor recovery. On the other hand, patients with moderate deficits spend significantly more time in a *weakly connected* state with reduced segregation, potentially picturing a signature of partially recovered motor function. In summary, employing a dynamic connectivity approach allowed for new insights into the systems-level effects that a stroke has on neural processing within the motor system, pointing to differential, reorganizational mechanisms depending on the initial motor deficit.

## Abbreviations

ARAT: action research arm test
dFNC: dynamic functional network connectivity
MRI: magnetic resonance imaging
BOLD signal: blood oxygenation level dependent signal
FDR: false discovery rate

## Acknowledgements

We are grateful to our colleagues at the Department of Neurology, University Hospital Cologne & Medical Faculty, University of Cologne for valuable support and discussions. Furthermore, we are grateful to our research participants without whom this work would not have been possible.

## Supplementary material

**Supplementary Table 1.** Demographics and clinical characteristics of stroke patient subgroups and healthy controls. Cortico-spinal tract (CST) affection was computed based on lesion overlap with a CST template provided in the SPM Anatomy Toolbox (Eickhoff *et al*., 2005).

|  | **Severely affected (n=13)** | **Moderately affected (n=18)** | **Healthy controls (n=17)** | **p-value** |
| --- | --- | --- | --- | --- |
| **Age (in years)** | 70.0 | 67.0 | 65.4 | Moderate-Controls: 0.65<br>Severe-Controls: 0.13 |
| **Sex (in % female)** | 61 | 22 | 12 | Moderate-Controls: 0.66<br>Severe-Controls: 0.01* |
| **Mean Framewise Displacement** | 0.29 | 0.27 | 0.27 | Moderate-Controls: 0.99<br>Severe-Controls: 0.59 |
| **NIHSS** | 10 | 6 | - | 0.01* |
| **ARAT *affected hand*** | 10 | 45 | - | 0.00* |
| **Lesion volume (in mm<sup>3</sup>, median)** | 12960 | 12732 | - | 0.61 |
| **CST overlap (in %, median)** | 9.2 | 4.8 | - | 0.68 |

